# Novel recruitment and retention strategies for sarcopenia trials: learning from the MET-PREVENT randomised controlled trial

**DOI:** 10.1101/2025.06.19.25329930

**Authors:** Claire McDonald, Nina Wilson, Katherine J Rennie, Michelle Bardgett, Penny Bradley, Andrew P Clegg, Stephen Connolly, Helen Hancock, Shaun Hiu, Karen Nicholson, Laura Robertson, Laura Simms, Alison J Steel, Claire J Steves, Bryony Storey, James M. S. Wason, Thomas von Zglinicki, Avan A Sayer, Miles D Witham

**Affiliations:** AGE Research Group, Translational and Clinical Research Institute, Faculty of Medical Sciences, Newcastle University, Newcastle upon Tyne, UK; NIHR Newcastle Biomedical Research Centre, Newcastle upon Tyne Hospitals NHS Foundation Trust, Cumbria, Northumberland, Tyne and Wear NHS Foundation Trust and Newcastle University, Newcastle upon Tyne, UK; Queen Elizabeth Hospital, Gateshead Health NHS Trust, Gateshead, NE9 6SX, UK; Biostatistics Research Group, Population Health Sciences Institute, Newcastle University, Newcastle upon Tyne, NE2 4AA, UK; Newcastle Clinical Trials Unit, Population Health Sciences Institute, Newcastle University, Newcastle upon Tyne, NE2 4AE, UK; Pharmacy Directorate, Newcastle upon Tyne Hospitals NHS Foundation Trust, Newcastle Upon Tyne NE7 7DN; Academic Unit for Ageing and Stroke Research, University of Leeds, Bradford Royal Infirmary, Bradford, BD9 6RJ, UK; Patient and Public Involvement Representative; Kings College London and St. Thomas’ Hospital, London, SE1 7EH, UK; Ageing Research Laboratories, Newcastle University Biosciences Institute, Newcastle upon Tyne, NE4 5PL, UK

**Author notes:** Correspondence to: Dr Claire McDonald, NIHR Newcastle Biomedical Research Centre, Newcastle University Campus for Ageing and Vitality, Newcastle upon Tyne, NE4 5PL, UK.

**Keywords:** Older People, Sarcopenia, Trials, Recruitment, Retention

## Abstract

**Background:** Recruiting and retaining participants in sarcopenia trials is challenging due to barriers in diagnosis, case-finding, exclusion criteria, frailty and drop-out. We describe and evaluate processes used by the MET-PREVENT randomised controlled trial to improve recruitment and retention.

**Methods:** MET-PREVENT was a two-centre, double-blinded, placebo-controlled trial examining the efficacy of metformin in patients with probable sarcopenia and either pre frailty or frailty. A telephone pre-screening step used the SARC-F questionnaire to identify those at risk of sarcopenia. The study was designed to maximise inclusion; and all study assessments could be conducted in participants homes or at a clinical research facility according to participant preference. Outcome measures were chosen to be simple and quick to collect with low burden to participants. Data on absolute numbers approached and randomised from a range of recruitment channels were analysed along with percentage conversion from approach to randomisation. The relationship between baseline factors at prescreening and conversion to randomisation (age, sex, use of walking aids, SARC-F score) was analysed to evaluated.

**Results:** 1630 people were approached and 268 people expressed interest in participation – of whom 214/268 (80%) underwent telephone pre-screening and 112/214 (42%) progressed to face-to-face screening.72/112 (64%) were randomised and 70/72 (97%) completed the trial. Recruitment took place from secondary care geriatric medicine clinics and via primary care mailshots; routes where patients had already had muscle strength recorded showed greater recruitment efficiency than those that did not. At face-to-face screening, SARC-F scores of 1 to 4 showed lower efficiency of recruitment compared to 5+ (49/82 [60%] vs 23/30 [77%] respectively) but accounted for most recruits (49/72 [68%]); age and sex were not associated with differences in recruitment. The majority (148/214 [69%]) of potential participants at prescreening expressed a preference for home visits; 101/112 (90%) undertook the screening visit at home and 45/72 (63%) of those randomised undertook either or both outcome visits at home.

**Conclusion:** A package of innovations in participant identification, recruitment processes and study visits enabled recruitment to target and the achievement of very high retention rates for a condition where it has traditionally been challenging to conduct clinical trials.

**Trial registration:** ISRCTN29932357

## Background

Sarcopenia is the age-related loss of muscle strength and mass [1]. It is a major cause of falls, prolonged hospital stay, diminished quality of life and earlier death in older people. It is associated with a significant socioeconomic burden due to impairment in activities of daily living and an increased risk of hospitalisation and admission to 24-hour care [2,3]. Resistance training is the only intervention with good evidence for the treatment of sarcopenia [4], but not all older people are willing or able to sustain the intensity of resistance training required. There are currently no approved pharmacological treatments for sarcopenia, and there is an unmet need for novel therapies.

Undertaking clinical trials for sarcopenia presents multiple challenges. Accurate diagnosis of the condition is not commonly made in clinical practice [5]. Even when the diagnosis is confirmed, it is often not systematically recorded in searchable electronic health records or databases. Consequently, identifying patients with sarcopenia for clinical trials is challenging. Furthermore, the target population for clinical trials in sarcopenia comprises older adults – a group who are generally underrepresented in clinical trials [6,7]. There are multiple underlying reasons for this lack of representation of older patients in clinical trials. Multimorbidity (the presence of two or more long-term conditions in an individual) is the norm for people living with sarcopenia [7,8], and the presence of both multiple conditions and the attendant polypharmacy often means older patients are underrepresented in trials based on restrictive exclusion criteria [9]. Intercurrent illnesses may also increase the risk of a participant withdrawing from the trial [10]. Older people with sarcopenia by definition have challenges with mobility and fatigue due to muscle weakness. Thus attending research appointments in distant research centres, and enduring long, complex study visits, pose additional barriers to participation. Successful trial design and delivery for people with sarcopenia therefore requires considering these potential barriers and changing trial design and delivery processes to better meet the needs of people living with sarcopenia if research results are to be representative and thus applicable to clinical practice [11].

MET-PREVENT was a phase II, randomised, double-blind, parallel-group, placebo-controlled, two-centre clinical trial examining the efficacy of metformin to improve physical performance in older patients with sarcopenia and either pre-frailty or frailty [12]. Here, we describe the approaches used to recruit and retain participants in the MET-PREVENT trial and evaluate the efficiency and effectiveness of aspects of the recruitment process.

## Methods

Full details of the MET-PREVENT trial protocol have been published previously [13] as have the main trial results [12]. MET-PREVENT included people aged 65 and over with probable sarcopenia [14] denoted by low handgrip strength (<16kg for women, <27kg for men) or prolonged five-times sit to stand time (>15s). Participants also had to have a 4m walk speed of <0.8m/s, thereby meeting the criteria for both sarcopenia and either prefrailty or frailty using the Fried phenotypic frailty criteria [15]. Participants were excluded if they had type 1 or type 2 diabetes mellitus, had previous intolerance to metformin or took metformin for another condition, had an estimated glomerular filtration rate of less than 45 mL/min/1.73 m^2^, liver impairment, were felt by the investigator to have a life expectancy of less than 3 months, or had a skeletal myopathy that was clearly due to an alternative cause. Participants were randomised to receive 4 months of metformin tablets 500mg three times per day or matching placebo. The primary outcome for the trial was the between-group difference in 4m walk speed at 4 months, adjusted for baseline values. A total of 72 participants were randomised to the trial, with 70 (97%) completing the four-month study visit. The trial was approved by the UK Health Research Authority North-West—Liverpool Central Research Ethics Committee (approval number 20/NW/0470) and by the UK Medicines and Healthcare products Regulatory Agency (trial reference number 2020-004023-16).

### Recruitment channels

Multiple recruitment channels were used to maximise recruitment opportunities. Two recruiting sites (Newcastle and Gateshead) took part in the trial, with each site using a different selection of methods. The recruitment channels used were:

#### Primary care

Electronic patient records (EPR) at a single primary care practice near Gateshead were electronically searched using automated searching, seeking patients aged 65 and over with an electronic frailty index score of 0.12 or greater [16], with exclusions for excessive alcohol consumption, abnormal liver function tests, chronic kidney disease stage 3b to 5, diagnosis of diabetes mellitus, use of metformin, or near end of life. Potential participants were invited to participate via mailshotsg.

#### Hospital clinics assessing sarcopenia

At Newcastle, participants were selected from a geriatric medicine clinic and from a geriatric medicine physiotherapy outpatients service that both measured grip strength and sit to stand test times. It was therefore possible to interrogate clinical records to support the identification of patients with probable sarcopenia who had previously attended these services, and patients without exclusion criteria on clinician notes review were then sent invitations.

#### Hospital clinics not assessing sarcopenia

Geriatric medicine clinics in Newcastle that did not assess grip strength or sit to stand were also included; those attending these clinics did so for a range of reasons including falls, declines in mobility, and follow-up after inpatient admission. Patients without exclusion criteria on clinician notes review were then sent invitations. Patients attending the osteoporosis service in Gateshead who were aged 75 and over were also invited.

#### Hospital acute frailty services

Patients presenting to the emergency department or medical assessment unit at Gateshead with frailty syndromes were assessed by the acute frailty team. Grip strength and sit to stand were not assessed, but patients with a Clinical Frailty Score [17] of between 4 (prefrailty) and 7 (severe frailty) were offered the chance to participate either by face-to-face approach or mailout to those who had attended over the preceding 12 months.

#### Community frailty service

In Newcastle, a community frailty service was used that measured grip strength and sit-to-stand test times, helping to identify patients with probable sarcopenia who had previously attended these services, and these patients were sent invitations to participate in the trial.

#### Specialist sarcopenia registry and previous sarcopenia trials

The SarcNet registry [18] was used to identify potential participants at the Gateshead site. SarcNet contains participants with probable sarcopenia willing to be contacted about future clinical studies, with 43 participants living in the Gateshead area. In addition, participants previously screened for the LACE multicentre randomised trial [19] in Newcastle were offered the opportunity to take part in MET-PREVENT.

#### General research registry

The NIHR Newcastle BioResource is a registry containing over 5000 participants of all ages who have consented for study recontact [20]. Participants joining since 2018 have completed the SARC-F score (a simple five-question screen for the possible presence of sarcopenia [21]). People aged 65 and over with a SARC-F score of 1 or more were sent information about MET-PREVENT.

#### Other

A small number of participants received invitations or expressed interest via routes outside these main channels, including relatives and friends of participants and those who heard about the trial via public talks or through other research studies.

### Initial Approach

People were invited to participate in the study following face-to-face contact with a clinician or via written invitation. Invitations were sent from a doctor in primary or secondary care via Docmail® (Docmail Ltd. Wells Rd, Radstock UK). Pre-paid envelopes in which to return expression of interest (EOI) forms directly to the research team were included. Where participants were approached face-to-face (e.g. in secondary care clinics), the approaching clinician could, with each participant’s consent, pass their details on to the research team. Potential participants were sent or given a brief study information sheet, EOI form and pre-paid envelope addressed to the research team to return the expression of interest form.

### Recruitment Pathway and Study visits

A prescreening telephone call was used to provide participants with an initial explanation of the study, ascertain the SARC-F score and establish the presence of any known exclusion criteria. Based on previous data, people with a SARC-F score of 1 or more were eligible to undergo a face-to-face screening visit [21]. People who passed pre-screening were offered a screening visit in their homes or at a local clinical research facility. Study visits and trial processes were designed to be as flexible as possible and to minimise burden to encourage retention in the trial. Key features of MET-PREVENT relevant to recruitment and retention are given in Box 1:

##### Key features of MET-PREVENT to facilitate recruitment and retention

###### Design

- Limited number of exclusion criteria
- Simple two step screening process with telephone pre-screen to minimise screening visits
- Regular but short study visits to stay in contact with participants
- All study outcomes could be conducted at participants own home
- Study outcomes chosen to minimise burden on participants
- Encouragement to remain in trial even if discontinued study intervention

###### Delivery

- All study visits could be conducted in participants home or in research centre
- Home visits conducted by investigators and research nurse teams with appropriate skills and experience working with older people
- Use of dedicated community research support team
- Regular participant newsletters

###### Public and patient involvement

- People at risk of or living with sarcopenia and frailty involved in design of trial, selection of outcomes, design of information, oversight of trial via trial management group, design and delivery of engagement events

## Data collection and analysis

As part of the data collection process for prescreening and screening visits, reasons for declining participation were collected, along with information on participant preference as to whether trial visits were conducted at home or in a research centre. Data were analysed using IBM SPSS® Version 29.0.1.0 (IBM, New York, USA). Data were graphed and visually inspected for normality. Continuous data are shown as mean and SD or median and IQR as appropriate. Normally distributed data from two groups were compared using Student’s t-test. Data that were not normally distributed were compared using the Mann–Whitney U test. Normally distributed data from three or more groups were compared using ANOVA. Categorical data are shown as number and percentage and compared using Pearson’s chi-squared test. A two-sided p value of <0.05 was taken as significant for all analyses

## Results

### Prescreening results

A total of 1630 individuals were invited to participate in the trial, and expressions of interest were received from 268 people; 214/268 people underwent telephone prescreening, 112/214 progressed to the screening visit and 72/112 were randomised between August 1^st^ 2021 and September 30^th^ 2022 as presented previously [13]. There was no evidence of a difference in age (80.3 [SD 5.8] vs 79.7 [SD 6.0] years, p=0.44) or sex (30 [42%] vs 71 [50%] men; p=0.25) between those who were randomised versus prescreened but not randomised. There was evidence of a difference on SARC-F score with those randomised having a higher median score (median 3 [IQR 2 to 5] vs median 2 [IQR 0 to 5]; p<0.001). Fifty-four people who expressed interest did not undergo telephone prescreening; reasons are shown in Supplementary Table 1. Reasons for non-eligibility at prescreening are given in Supplementary Table 2.

### Participant characteristics at prescreening by recruitment route

Table 1 shows the characteristics of those expressing interest in the study by recruitment channel. Participants recruited via research registries or community frailty services were on average younger than those recruited via secondary care or primary care clinics. The SARC-F score was on average lower for individuals who responded to an invitation sent through primary care or the general research registry, compared to those recruited through secondary care, the specialist sarcopenia registry or the acute frailty service had the highest SARC-F score. Consistent with this finding, those recruited through primary care or general research registries were also less likely to use a walking aid.

**Table 1.**
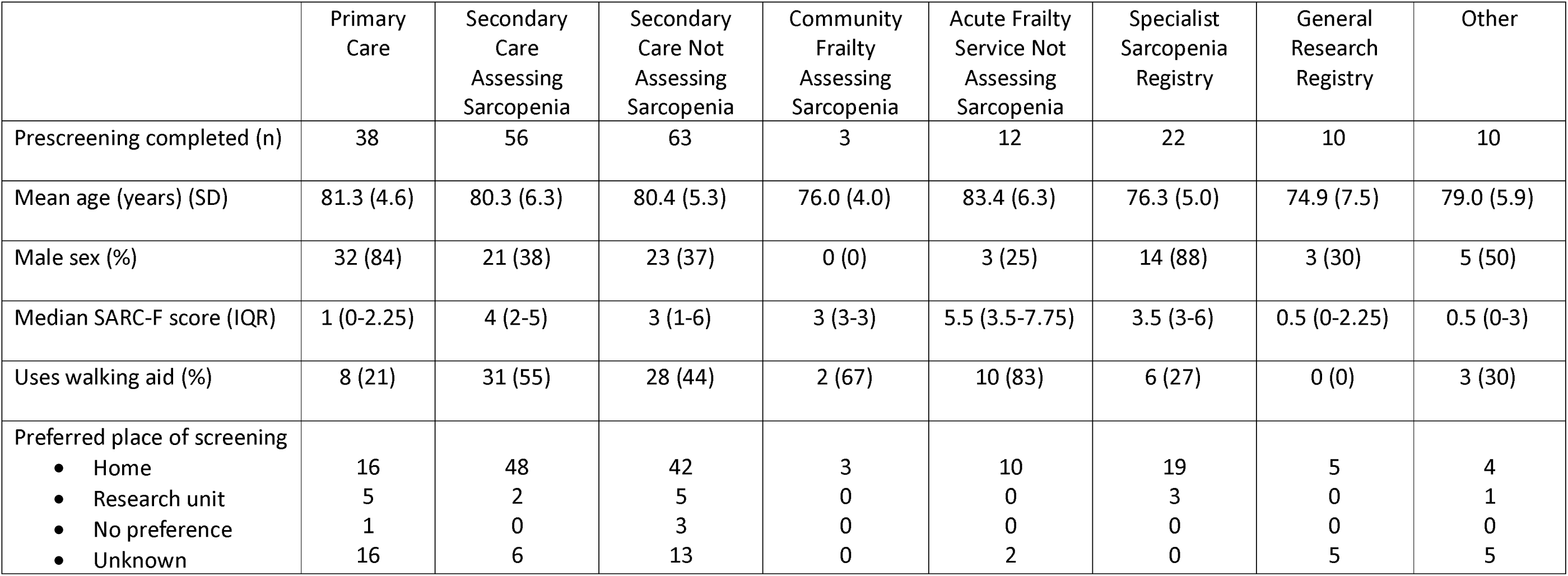
Characteristics of people undergoing prescreening from different recruitment channels in MET-PREVENT trial.

|  | Primary Care | Secondary Care Assessing Sarcopenia | Secondary Care Not Assessing Sarcopenia | Community Frailty Assessing Sarcopenia | Acute Frailty Service Not Assessing Sarcopenia | Specialist Sarcopenia Registry | General Research Registry | Other |
| --- | --- | --- | --- | --- | --- | --- | --- | --- |
| Prescreening completed (n) | 38 | 56 | 63 | 3 | 12 | 22 | 10 | 10 |
| Mean age (years) (SD) | 81.3 (4.6) | 80.3 (6.3) | 80.4 (5.3) | 76.0 (4.0) | 83.4 (6.3) | 76.3 (5.0) | 74.9 (7.5) | 79.0 (5.9) |
| Male sex (%) | 32 (84) | 21 (38) | 23 (37) | 0 (0) | 3 (25) | 14 (88) | 3 (30) | 5 (50) |
| Median SARC-F score (IQR) | 1 (0-2.25) | 4 (2-5) | 3 (1-6) | 3 (3-3) | 5.5 (3.5-7.75) | 3.5 (3-6) | 0.5 (0-2.25) | 0.5 (0-3) |
| Uses walking aid (%) | 8 (21) | 31 (55) | 28 (44) | 2 (67) | 10 (83) | 6 (27) | 0 (0) | 3 (30) |
| Preferred place of screening |  |  |  |  |  |  |  |  |
| • Home | 16 | 48 | 42 | 3 | 10 | 19 | 5 | 4 |
| • Research unit | 5 | 2 | 5 | 0 | 0 | 3 | 0 | 1 |
| • No preference | 1 | 0 | 3 | 0 | 0 | 0 | 0 | 0 |
| • Unknown | 16 | 6 | 13 | 0 | 2 | 0 | 5 | 5 |

### Efficiency and effectiveness of different recruitment routes

Table 2 shows the efficiency of each recruitment channel (i.e. the proportion of people approached who were randomised into the trial) and the effectiveness of each recruitment channel (the absolute number of recruits for each channel). The efficiency of recruitment was highest for channels that had already measured muscle strength (some secondary care clinics, specialist registries and the community frailty service); efficiency was higher for secondary care services that did measure muscle strength compared to those that did not (38/316 [12%] vs 17/505 [3%]; p<0.001). Effectiveness (overall volume of recruits) was highest from secondary care clinics, with primary care also contributing several participants despite a relatively small number of primary care invitations being sent.

**Table 2.** Efficiency and effectiveness of different recruitment channels in MET-PREVENT trial.

|  | Primary Care | Secondary Care Assessing Sarcopenia | Secondary Care Not Assessing Sarcopenia | Community Frailty Assessing Sarcopenia | Acute Frailty Service Not Assessing Sarcopenia | Specialist Sarcopenia Registry | General Research Registry | Other |
| --- | --- | --- | --- | --- | --- | --- | --- | --- |
| Invited (n) | 200 | 316 | 505 | 34 | 345 | 37 | 170 | 23 |
| Pre-screening (n) | 38 | 56 | 63 | 3 | 12 | 22 | 10 | 10 |
| Attended screening visit (n) | 15 | 40 | 29 | 3 | 3 | 14 | 4 | 4 |
| Randomised (n) | 9 | 38 | 17 | 3 | 1 | 2 | 2 | 0 |
| Conversion from invitation to randomisation (%) | 5 | 12 | 3 | 9 | <1 | 10 | 1 | 0 |

### Prescreening characteristics as predictors of progression to randomisation

Supplementary Table 3 shows prescreening characteristics as predictors of progression to randomisation. There was no evidence that age and sex were associated with differences in progression to randomisation, but use of a walking aid was associated with progression to randomisation (42/88 (47.7%) vs 30/126; (23.8%) p<0.001). Eligibility at the screening visit and conversion rate to randomisation by SARC-F score is shown in Figure 1. SARC-F scores above 4 were associated with a non-statistically significant higher recruitment efficiency (i.e. higher percentage conversion from screening to randomisation) (23/30 [77%] vs 49/82 [60%]; p=0.10) but the majority of participants randomised (49/72 [68%]) had a SARC-F score between 1 and 4.

**Figure 1a.**
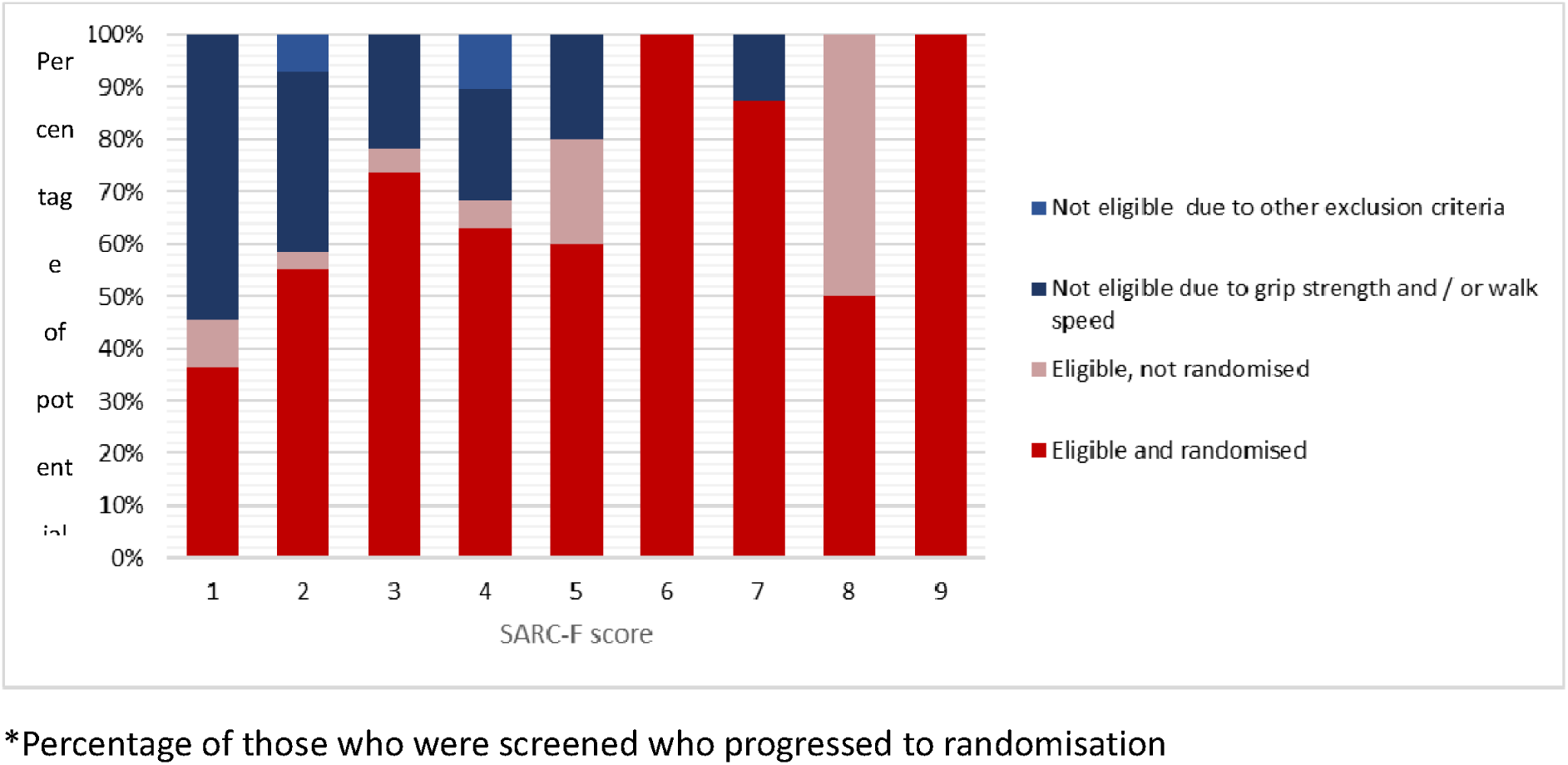
Efficiency* of participant selection at screening visit for different SARC-F scores. *Percentage of those who were screened who progressed to randomisation

**Figure 1b.**
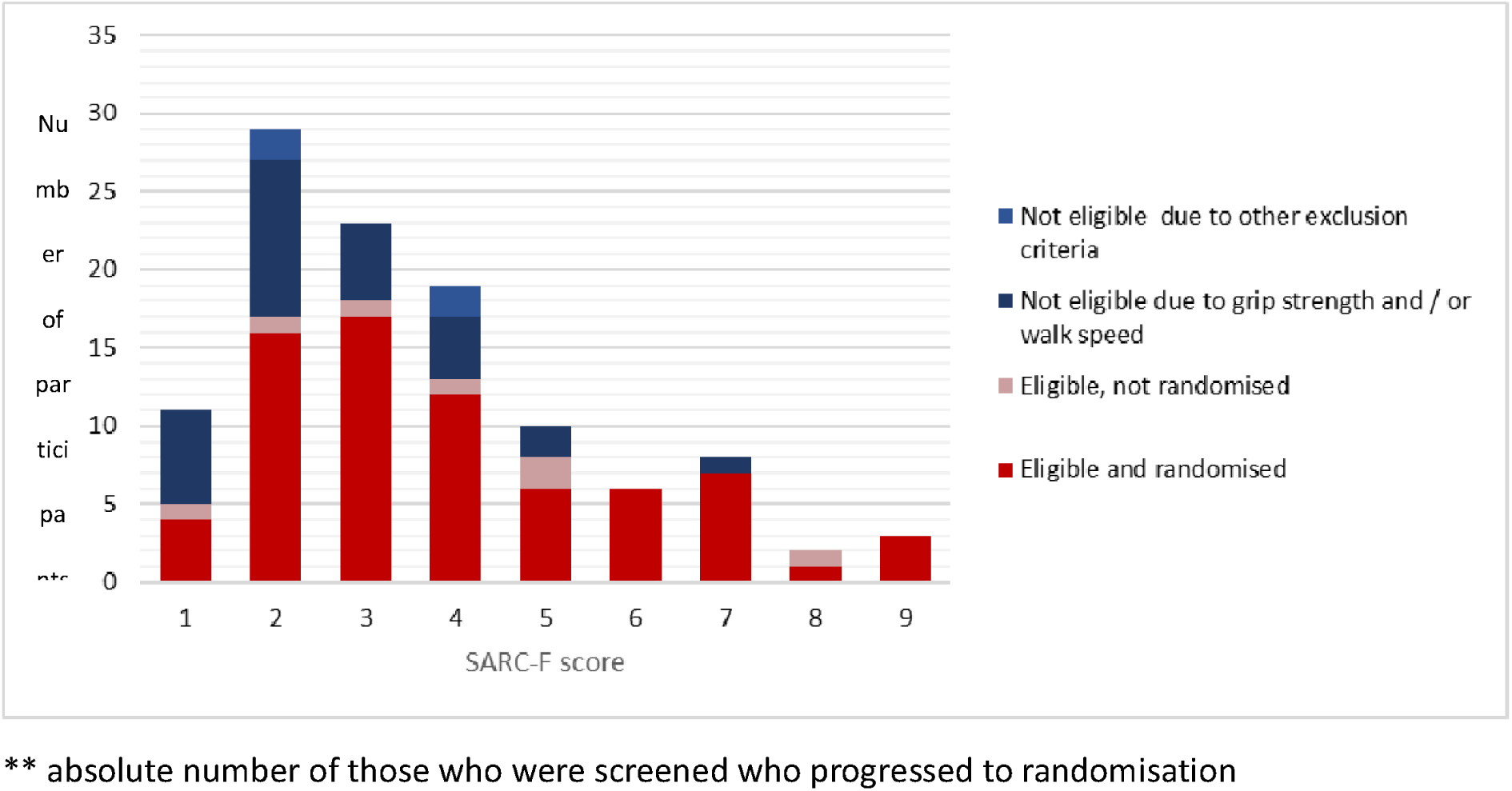
Effectiveness** of participant selection at screening visit for different SARC-F scores. ** absolute number of those who were screened who progressed to randomisation

### Home visits

Most participants completing prescreening (137/198 [69%]) stated that their preferred place of screening visit was at home. Participants opting for a home visit (n=137) did not differ in age compared to those who preferred to attend clinic (n=61) [mean age 80.2 (SD 5.9) vs. 79.5 (SD 5.8); p=0.33] but did have significantly higher SARC-F scores (median 4 [IQR 2 to 5) vs 0 [IQR 0 to 3]; p<0.001]. Women were more likely to choose a home visit than men (92/113 [81%] vs 55/101 [54%]; p<0.001], as were participants using a walking aid (72/88 [81%] vs 75/126 [60%]; p <0.001] when asked at the prescreening telephone call. 101/112 (90%) of participants underwent the screening visit at home, and 45/72 (63%) of participants who were randomised underwent either or both the baseline visit and the follow-up visit at home.

## Discussion

We describe a package of design and delivery features from the MET-PREVENT sarcopenia and frailty trial that helped to successfully recruit and retain participants in a field where conducting clinical trials has traditionally been challenging. Key features included minimising exclusion criteria and trial visit burden, using a telephone prescreening call and a simple questionnaire to identify participants likely to be eligible despite no formal diagnosis of sarcopenia, and a flexible approach to study delivery with options for delivering study visits in participants own homes. We found that secondary care geriatric medicine clinics, and primary care, were the major sources of recruitment in terms of numbers, that clinics where muscle strength was already measured were more likely to yield expressions of interest that translated into randomised recruits, and that a higher cutoff for the SARC-F score used in prescreening was associated with higher rates of conversion to randomisation but at the cost of missing many people who would be eligible to take part in such trials.

Older adults are willing to take part in research but have historically been prevented from doing so by overt and covert ageism and exclusion criteria [23,24]. Older adults often consider specific study design characteristics when deciding whether to participate in a trial. These factors include the aim of the intervention, medication issues, adverse events, but also practical issues such as the frequency and duration of study visits, caregiver responsibility, use of new technology, difficulty getting to research sites, transportation problems and mobility issues [6, 25,26]. Current guidance promotes the need for inclusive, flexible designs that overcome these barriers [11], along with the need to co-design trials with older people. We followed this guidance in designing and delivering MET-PREVENT and the high number of participants expressing a preference for and taking up the offer of home visits suggests that this flexible approach to delivery is key to success.

Home visits in clinical trial delivery offer several advantages, particularly in improving patient recruitment, reducing missed visits and missing data—as demonstrated in the MET-PREVENT trial— and enhancing participant retention. Many participants prefer these visits, which can lead to better engagement and compliance throughout the study. However, there are important considerations, especially concerning cost and logistics. While sending research staff to participants’ homes incurs time and travel expenses, it can be more cost-effective than arranging transportation for patients to attend clinic visits, especially for populations with mobility issues. Additionally, utilising home visits can reduce the reliance on expensive clinical space. Despite these benefits, not all tests can be conducted in community settings and some participants’ home environments may not be suitable for certain procedures. Not all organizations will fund or support home visit models, and some research teams may be reluctant or unprepared to work in community settings. Crucially, the costs associated with home visits must be carefully and realistically included in funding applications, as these expenses can be easily underestimated. Balancing these factors is essential when determining whether home visits are appropriate for a particular clinical trial [27]

Recruitment to sarcopenia trials has traditionally been challenging with trials struggling to recruit to target despite sarcopenia affecting 5-10% of adults. Absolute numbers recruited are the primary consideration for any trial, but to maximise the time and effort spent recruiting, recruitment needs to be as efficient as possible – i.e. it should yield the most participants enrolled for the least effort. The conversion rates we found from invitation to randomisation are considerably higher than in some previous sarcopenia trials, where conversion rates have been around 1% [28], and the rate of conversion from screening visit to randomisation was higher for MET-PREVENT than for previous trials which found conversion rates of between 10 and 50% [28–30]. This was in part due to the availability of muscle strength measures for patients recruited from some clinics and services, suggesting that efforts to embed muscle strength measurement into clinical practice should be an important component of readying sites to recruit to sarcopenia trials.

Research registries have been proposed to improve recruitment to clinical research [31]. Despite both the sarcopenia specific registry and the general registry recording SARC-F, the sarcopenia-specific registry performed better than the general registry in terms of response rate and overall efficiency. A final observation from different recruitment channels is that although the conversion rate from primary care was lower than from some secondary care clinics, the size of the primary care population that can be reached by mailshots is far larger, making primary care recruitment an attractive way to deliver the required volume of participants despite lower efficiency. Similar results have been seen in previous sarcopenia trials [28].

The SARC-F score is a simple set of questions that can easily be used to select potential participants at prescreening, either in person, by telephone or by web-based methods. The original cutoff for identifying patients likely to have sarcopenia was proposed to be 4 out of 10, however, it is clear that such a cutoff is specific but not sensitive for detecting sarcopenia [32]. Previous work has suggested that a lower cutoff of 1 or more points could identify a greater number of people with sarcopenia [22], and the results from our analysis support this approach. Greater efficiency (due to higher specificity) was seen with higher SARC-F scores, but the majority of randomised participants had SARC-F scores between 1 and 4. Even at this lower level, conversion rates from prescreening to randomisation were 50% or higher suggesting that a cutoff of 1 provides an acceptable balance of efficiency and volume for participant selection.

Our analysis has a number of strengths. Few sarcopenia trials have attempted to collect systematic information on recruitment processes, and not all sarcopenia trials have been co-designed with patient involvement to provide a package of features to enhance recruitment and retention. Several limitations should also be acknowledged, however. We were unable to collect fully comprehensive data on all parts of the recruitment process. Participants did not always give reasons for choosing not to proceed with the study, and reasons given in brief responses may not reflect deeper motivations or barriers. We were unable to collect data on all participants who received an invitation (particularly from primary care) due to data protection considerations. We did not test components of our package of design and delivery features in a randomised trial (e.g. a Study Within a Trial; SWAT), thus caution is needed in attributing causality. There is more to do in terms of further developing trial design and processes to enhance inclusivity, for example to better engage with ethnically minoritised groups, those with low literacy levels and those with visual or hearing impairments. Finally, our trial recruited during and just after the second wave of the COVID-19 pandemic, and circumstances and expectations around healthcare delivery during that period may not reflect current and future context for delivering trials.

### Implications for practice

We have shown that is it possible to design clinical trials that can successfully recruit and retain older people with sarcopenia and pre-frailty or frailty with a much better retention rate than has been achieved in previous similar trials. Similar principles could be employed in future sarcopenia trials; our findings also strongly suggest that sites should prepare for recruitment by implementing routine measurement and systematic recording of muscle strength (grip strength and sit to stand tests) into routine clinical practice to enable participant identification; sarcopenia-specific registries could also help identify participants. Research delivery teams need to ensure that they have the skills and processes in place to support trial visits at home, and trial protocols need to allow such visits. Our results will be helpful to research teams seeking to design and deliver trials that meet the needs of older people with sarcopenia and frailty, enabling recruitment to time and target, enrolling older people who are truly representative of those seen in clinical practice, with high retention to minimise bias from drop-out.

## Data Availability

De-identified individual participant-level data from this trial are stored at the NIHR Newcastle Biomedical Research Centre and will be made available to other researchers subject to completion of appropriate collaboration agreements and data sharing agreements with the trial team. For access to trial documents (information sheet, consent form, protocol, statistical analysis plan), contact the chief investigator.

## Acknowledgements

CM, AAS and MDW acknowledge support from the NIHR Newcastle Biomedical Research Centre and the Multiple Long-term Conditions Cross-NIHR Collaboration (MLTC CNC). MDW acknowledges support from the NIHR HealthTech Research Centre in Diagnostic and Technology Evaluation and the NIHR Newcastle Clinical Research Facility. AC and JW are funded by NIHR Research Professorship awards; AC is also supported by the NIHR Applied Research Collaboration Yorkshire & Humber, the NIHR Leeds Biomedical Research Centre, and Health Data Research UK, an initiative funded by UK Research and Innovation Councils, NIHR and the UK devolved administrations and leading medical research charities. The views expressed in this publication are those of the author(s) and not necessarily those of the NIHR, NHS or the UK Department of Health and Social Care.

The MET-PREVENT trial was delivered with the help of the NIHR North-East and North Cumbria Regional Research Delivery Network and was run through the NIHR Newcastle Clinical Research Facility.

## Authors Contributions

MDW, APC, HH, CM, AAS, CJS, TvZ, and JMSW designed the trial. MDW, CM, MB, PB, SC, KN, KJR, LR,

AJS, BS, and LS conducted the trial. NW and SH accessed and verified the underlying study data with oversight from JW. CM and MDW conducted the analysis under the supervision of NW and drafted the manuscript. All authors had access to the data, interpreted the results, critically revised the manuscript, and all authors approved the final version for publication.

## Ethics statement

The trial was approved by the UK Health Research Authority North-West—Liverpool Central Research Ethics Committee (approval number 20/NW/0470). The trial is registered on the ISRCTN trial database (ISRCTN29932357). The trial was conducted according to the principles of the 1964 Declaration of Helsinki and its later amendments. Written informed consent was obtained from all participants.

---

**Supplementary Figure 1.**
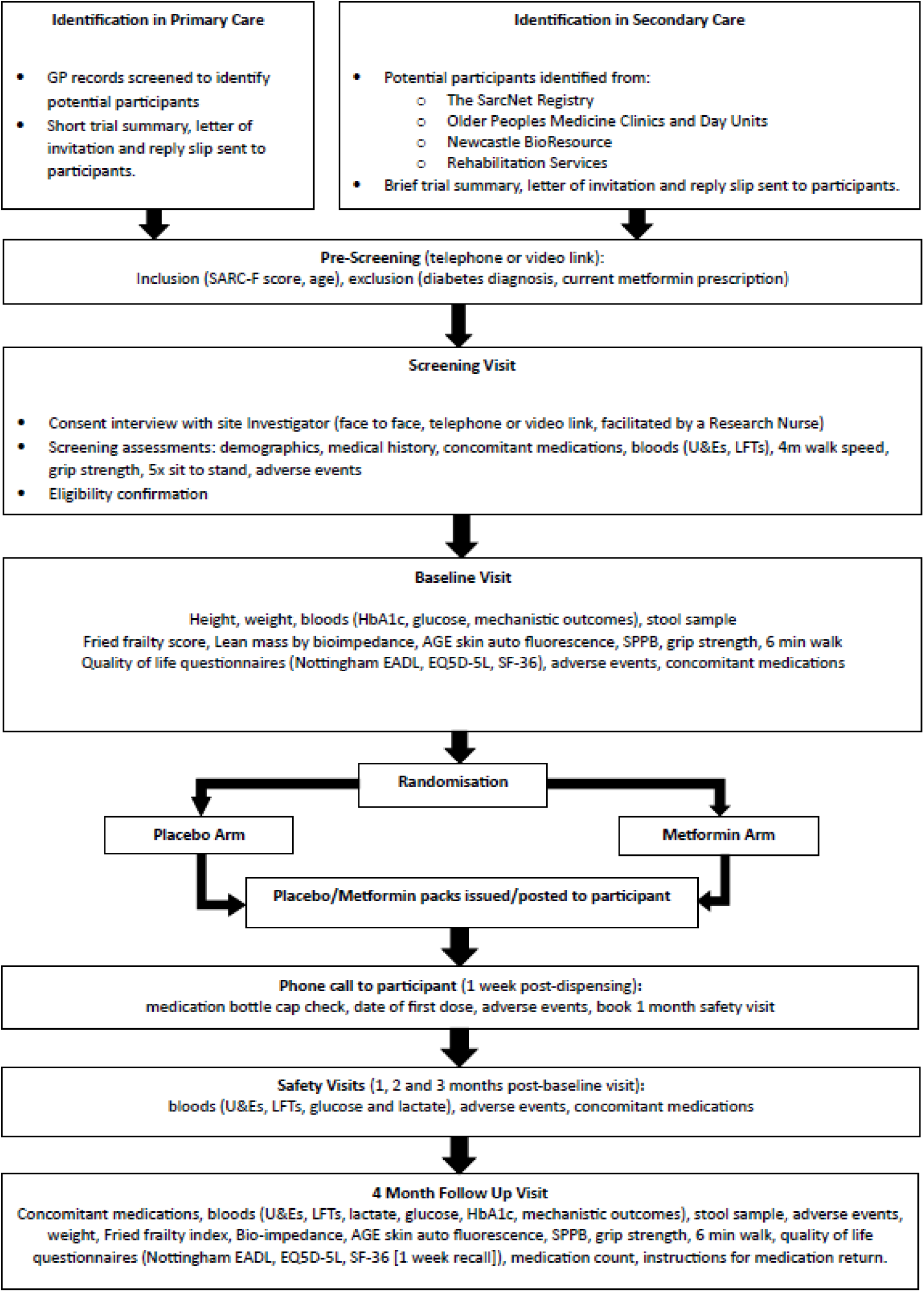
Recruitment process and study visits for the MET-PREVENT trial

**Supplementary Table 1.** Reasons for non-completion of prescreening call (n=54)

|  |  |
| --- | --- |
| Trial too burdensome | 5 |
| Ill health | 4 |
| Felt too old to take part | 1 |
| Felt too fit to take part | 3 |
| Did not want to take metformin | 3 |
| One or more exclusion criteria reported | 9 |
| Did not appear to understand study sufficiently to continue conversation | 9 |
| No reason given | 15 |
| No response to multiple telephone calls | 2 |
| No reason recorded | 3 |

**Supplementary Table 2.** Reasons for ineligibility at prescreening call (n=102)

|  |  |
| --- | --- |
| SARC-F score of 0 | 39 |
| Exclusion criteria on notes review | 2 |
| Unlikely to have capacity to consent on notes review | 2 |
| Declined to participate after reading full Participant Information Sheet: |  |
| Trial too burdensome | 3 |
| Ill health | 1 |
| Felt too fit | 1 |
| Did not wish to take metformin | 3 |
| Concerns from family | 1 |
| No reason given | 20 |
| Screening visit cancelled by trial team due to COVID pandemic | 3 |
| Screening visit not offered as trial recruitment target met | 27 |

**Supplementary Table 3.** Prescreening predictors of progression to randomisation.

| Characteristic |  | Number progressing to randomisation /<br>number at prescreening (%) | P* |
| --- | --- | --- | --- |
| Age | 65-74 | 14/33 (42) | 0.22 |
|  | 75-84 | 40/136 (29) |  |
|  | 85+ | 18/45 (40) |  |
| Sex | Men | 30/101 (30) | 0.31 |
|  | Women | 42/113 (37) |  |
| Use of walking aid | Yes | 42/88 (48) | <0.001 |
|  | No | 30/126 (24) |  |
| SARC-F score** | 1 | 4/18 (22) | 0.27 |
|  | 2 | 16/36 (44) |  |
|  | 3 | 17/32 (53) |  |
|  | 4 | 12/27 (44) |  |
|  | 5 | 6/20 (30) |  |
|  | 6 | 6/18 (33) |  |
|  | 7 | 7/12 (58) |  |
|  | 8 | 1/6 (17) |  |
|  | 9 | 3/5 (60) |  |

\*Using Pearson's Chi-squared test \*\*Data for SARC-F = 0 omitted as these participants were excluded from progressing to screening visit as per the protocol

